# Equity-Enhanced Glaucoma Progression Prediction from OCT with Knowledge Distillation

**DOI:** 10.1101/2024.12.31.24319264

**Authors:** Sulaiman O. Afolabi, Leila Gheisi, Jing Shan, Lucy Q. Shen, Mengyu Wang, Min Shi

## Abstract

**Purpose:** To develop an equitable deep learning model with knowledge distillation to enhance the demographic equity in glaucoma progression prediction.

**Methods:** We developed a novel deep learning model called FairDist which used baseline optical coherency tomography (OCT) scans to predict glaucoma progression. First, an equity-aware EfficientNet termed EqEffNet was trained for glaucoma detection. Next, the pretrained detection model was adapted for progression prediction using knowledge distillation which minimizes image and identity feature differences between the detection and progression models. Progression was defined based on longitudinal visual field maps from at least five visits up to six years. Model performance was measured by the area under the receiver operating characteristic curve (AUC), Sensitivity, Specificity, and equity was assessed using equity-scaled AUC (ES-AUC), which adjusts AUC by accounting for subgroup disparities, focusing on gender and racial groups.

**Results:** Two types of glaucoma progression including mean deviation (MD) Fast progression and Total Deviation (TD) Pointwise progression were explored. For MD Fast Progression, FairDist achieved the highest AUC and ES-AUC for gender (AUC: 0.738, ES-AUC: 0.693) and race (AUC: 0.778, ES-AUC: 0.677) compared with methods with and without integrating inequity mitigation strategies. For TD Pointwise progression, FairDist achieved the best AUC and ES-AUC for gender (AUC: 0.743, ES-AUC: 0.719) and race (AUC: 0.746, ES-AUC: 0.645) among all methods.

**Conclusions:** FairDist enhances both model performance and equity in glaucoma progression prediction after integrating the equity-aware learning and knowledge distillation components. The proposed deep learning model shows promise in improving glaucoma diagnosis while reducing disparities across demographic groups.

## Introduction

Glaucoma is the second-leading cause of blindness worldwide characterized by progressive vision loss (**Figure 1a**).^1^ It impacts over 3 million patients in the US and projections indicate that by 2050 this number will escalate to 6.3 million due to aging population, with African Americans and Hispanics being disproportionately affected.^2^ The irreversible nature of visual impairment from glaucoma progression underscores the importance of detecting visual dysfunction in its early stages.^3^ Glaucoma progression has traditionally been assessed by tracking the decline in visual function sensitivity over time using standard automated perimetry, though this approach is susceptible to substantial test-retest variability and confounding effect of age-related changes.^4^ Current standard clinical care for glaucoma also includes optical coherence tomography (OCT) imaging, which allows for the detection of progressive morphological changes, such as retinal nerve fiber layer (RNFL) thinning, closely associated with vision loss.^5^ Recent studies have developed deep learning-based methods to detect visual field (VF) deterioration from longitudinal OCT scans and retinal nerve fiber layer (RNFL) thickness, achieving promising area under the ROC curve (AUC) scores.^6,7^ However, OCT measurements over an extended period, often spanning several years, are typically needed to identify progression status,^8^ which makes it challenging to collect sufficient data for training a generalizable progression prediction model. In addition, methods that rely on RNFL thickness may be affected by OCT layer segmentation artifacts.^9,10^ In contrast, we develop a deep learning model which used the baseline OCT scans to predict glaucoma progression status. The glaucoma progression was defined by longitudinal VF data from at least five consecutive visits over a six-year period (**Figure 1b**).

**Figure 1:**
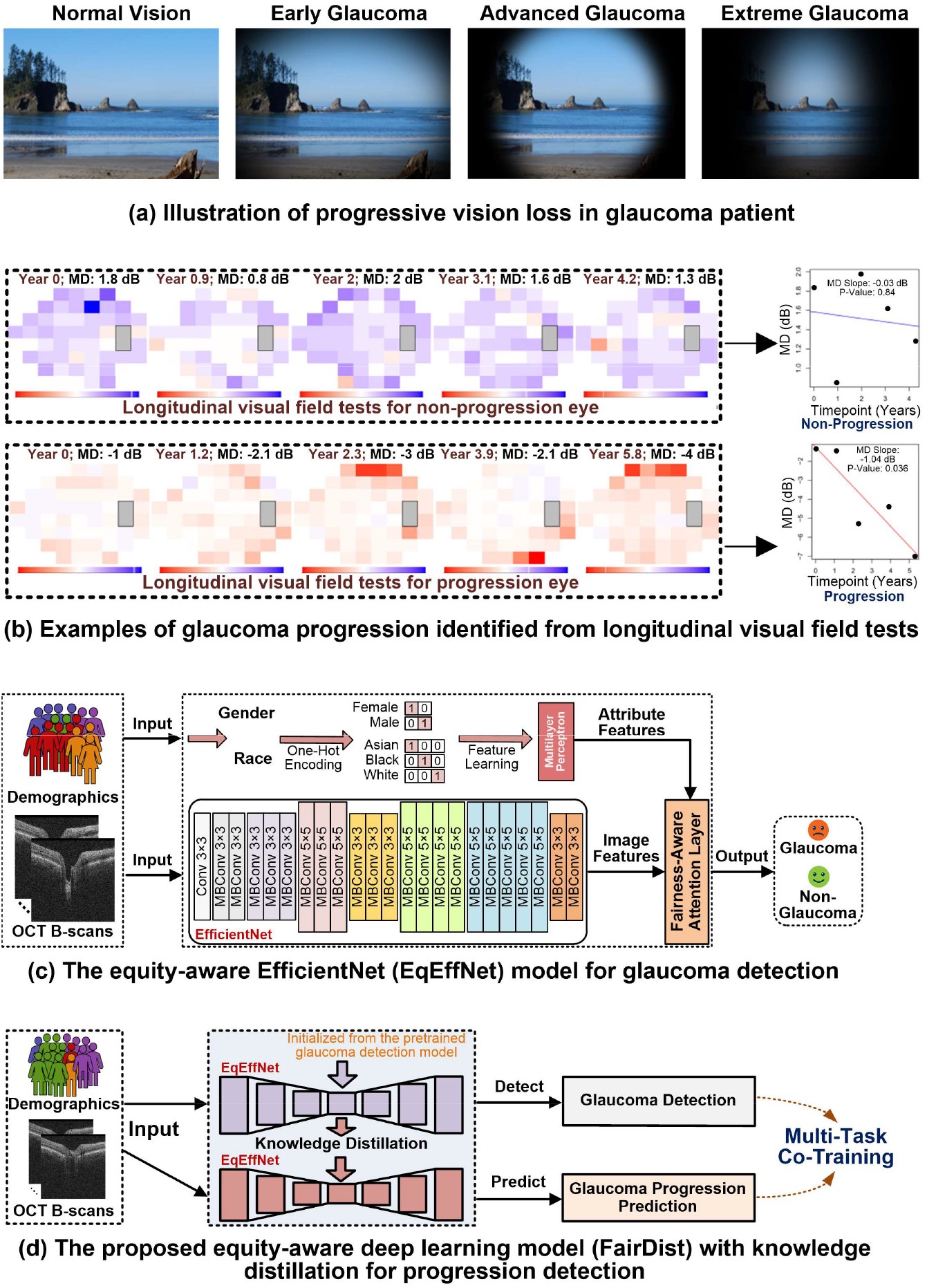
Illustration of glaucoma progression and the proposed equity-aware deep learning model. (a) Progressive vision loss in glaucoma patient. (b) Glaucoma progression is defined by longitudinal visual field data. (c) The proposed equity-aware EfficientNet, called EqEffNet, for training a glaucoma detection model. (d) The proposed framework, called FairDist, which generalizes the pretrained glaucoma detection model to enhance the demographic equity in glaucoma progression prediction through knowledge distillation.

On the other hand, while deep learning has the potential to improve the efficiency and accessibility of glaucoma diagnosis for a broader population,^11–14^ their performance across different demographic groups is underexplored. A growing body of research indicates that deep learning algorithms can reinforce and amplify biases present in the data,^15–19^ raising significant fairness concerns, particularly for certain racial and ethnic minorities. For example, a recent study found that Black individuals experience significantly lower performance outcomes compared to Whites and Asians when using RNFL thickness and OCT scans for glaucoma detection.^18^ Two other studies offered a comprehensive analysis of inequity concerns in artificial intelligence (AI) across various healthcare applications,^17,20^ especially impacting the underrepresented communities. The unfairness of AI models can stem from several factors. First, skewed data distributions in diagnosis labels and sensitive attributes may result in inadequate feature representation for specific groups. Second, anatomical differences between subgroups can create varying levels of difficulty for AI models to provide accurate diagnoses. Third, annotation inconsistencies arising from clinicians’ varying preferences based on patient attributes can confuse AI models and contribute to bias. Last, the AI model itself can perpetuate unfairness. Conventional AI training processes prioritize maximizing overall performance, which may inadvertently widen performance gaps between subgroups. Regardless of different reasons, it is of paramount importance to explicitly address the potential unfairness of AI models for equitable medical diagnosis.

In this work, we propose an equity-aware deep learning model to address these limitations. From the analyses above, two obstacles may hinder the development of a responsible progression prediction model: 1) the limited availability of longitudinal data with annotations and 2) potential disparities in prediction performance across different demographic groups. To address the challenge of limited data, we proposed training a glaucoma detection model (**Figure 1c**) using a large dataset and subsequently generalizing it for progression prediction through a knowledge distillation process (**Figure 1d**). Additionally, we introduced an equity-aware feature learning mechanism which adjusts the importance of OCT scan features based on patient identity information. The proposed integration of knowledge distillation and equitable feature learning aims to enhance both the performance and fairness of deep learning models for glaucoma progression prediction. The terms equity and fairness are used interchangeably in this paper.

## Methods

Two datasets were used to develop deep learning models for glaucoma progression prediction. This study complied with the guidelines outlined in the Declaration of Helsinki. In light of the study’s retrospective design, the requirement for informed consent was waived.

### Dataset Description

#### Glaucoma Detection Dataset

It is a large-scale glaucoma detection dataset to develop the glaucoma detection model. The dataset contains 10,000 reliable (signal strength is greater than 6) spectral-domain OCT (Cirrus, Carl Zeiss Meditec, Dublin, California) samples of 10,000 patients from the Massachusetts Eye and Ear glaucoma service between 2010 and 2022. The average age of patients was 60.7 ± 6.3 years. Each 3D OCT data sample contains 200 B-scans, each with a dimension of 200 ×200. The self-reported patient demographic information was as follows (**Figure 2a**): 57.0% of the patients are female and 43.0% are male; racially, 8.5% are Asian, 14.9% are Black, and 76.6% are White. Glaucomatous status was defined based on a reliable Humphrey VF test, which included a fixation loss of ≤ 33%, false-positive rate of ≤ 20%, and false-negative rate of ≤ 20%. A VF test conducted within 30 days of OCT was utilized to identify glaucoma patients. Criteria for the present of glaucoma include a VF mean deviation less than -3 dB, coupled with abnormal results on both the glaucoma hemifield test and the pattern standard deviation.^21^ Accordingly, 51.3% of the patients are categorized as non-glaucoma and 48.7% as glaucoma.

**Figure 2:**
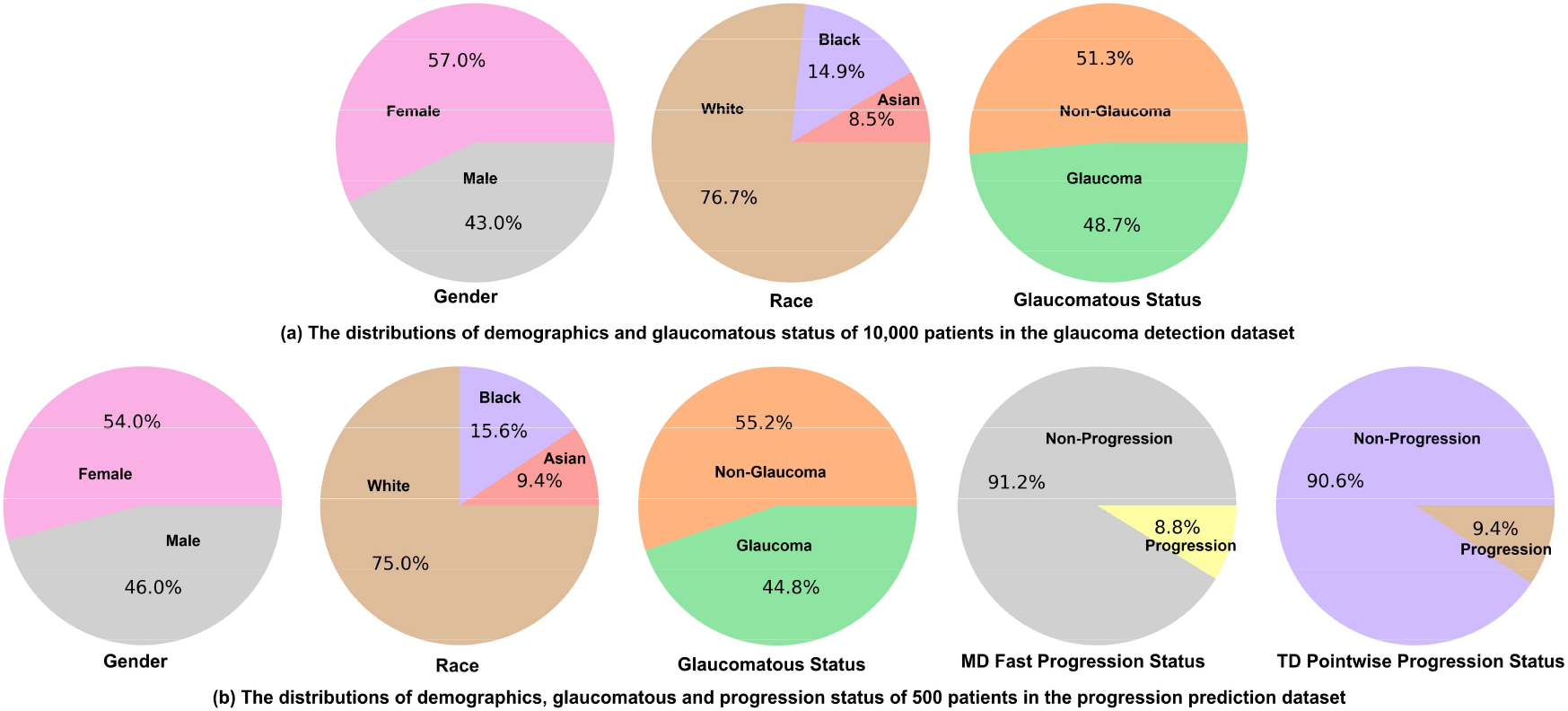
Distributions of demographics and labels in the adopted datasets. (a) Glaucoma detection dataset. (b) Glaucoma progression prediction dataset.

#### Glaucoma Progression Prediction Dataset

It is a public dataset for both glaucoma detection and progression prediction tasks. It includes 500 spectral-domain Cirrus OCT samples of 500 glaucoma patients from the Massachusetts Eye and Ear. The average age of patients was 62.9 ± 6.4 years. Each OCT sample contains 200 B-scans accompanying both glaucoma detection and progression prediction labels. Demographic information was as follows: Gender distribution is 54.0% female and 46.0% male; racial composition includes 9.4% Asian, 15.6% Black, and 75.0% White. The patients were divided into non-glaucoma and glaucoma categories, making up 55.2% and 44.8% of the dataset, respectively. The definition of glaucomatous status used the same criteria as in above glaucoma detection dataset. Two criteria were used to assess glaucoma progression based on VF maps collected over a minimum of five visits within a six-year period,^22^ with each map represented by a vector of 52 total deviation (TD) values ranging from -38 dB to 26 dB. The criteria are: (1) MD Fast Progression: eyes with an MD slope ≤ -1 dB. (2) TD Progression: eyes with at least three locations exhibiting a TD slope ≤ -1 dB; Note that the 500 samples included are baseline OCT scans and the VF data used to define the labels are private. For MD fast progression, 91.2% are categorized as non-progression and 8.8% as progression.^21^ For TD progression, the percentages are 90.6% for non-progression and 9.4% for progression.

#### The Proposed Equity-Aware Deep Learning Model with Knowledge Distillation

We developed an equity-aware deep learning model called EqEffNet for glaucoma detection (**Figure 1c**), which was afterwards generalized to a glaucoma progression prediction model called FairDist through the knowledge distillation (**Figure 1d**). EqEffNet used EfficientNet as the backbone model and integrated a fair attention mechanism to extract features from the OCT scans. The EfficientNet architecture comprises seven blocks differentiated by color, each consisting of multiple MBConv layers, a mobile inverted bottleneck convolution design that integrates squeeze-and-excitation optimization. These blocks vary in the depth and size of their filters as indicated by the kernel sizes and the number of layers in each block. Starting with the initial standard convolution layer, the network progresses through these MBConv blocks, which increase in complexity and depth. Each block represents a stage in feature extraction where similar operations are applied to the input features. This hierarchical structure allows EfficientNet to efficiently manage computational resources while maximizing learning and representational capacity, making it highly effective for tasks requiring detailed image analysis and classification. However, EfficientNet may exhibit biases towards certain demographic groups (e.g., Asians and Blacks), resulting in notable performance discrepancies among different populations. To address this, we introduced a fairness-aware attention mechanism that adjusts feature learning based on demographic attributes. Specifically, we utilized a multilayer perceptron encoder to process the binary-encoded demographic attributes associated with the input scans. Afterwards, the learned attribute and image features were used to calculate the importance weights for each of image features. Therefore, image features could adjust their importances for identity attributes to obtain balanced attentions and contributions for the final glaucoma detection and progression prediction outcomes. The glaucoma detection model EqEffNet was trained using the Glaucoma Detection Dataset including 10,000 OCT samples. 60%, 10% and 30% of the samples were used for model training, evaluation and testing, respectively.

Next, the pretrained glaucoma detection model was used to initialize a glaucoma detection model which guided the training of a glaucoma progression prediction model termed FairDist (**Figure 1d**). Both glaucoma detection and progression prediction models used EqEffNet to learn features, and these two EqEffNet models took the same OCT scans as inputs but performing glaucoma detection and progression prediction tasks, respectively. At the same time, the pretrained glaucoma detection model (as a teacher) empowered the progression prediction model (as a student) through a knowledge distillation process which minimizes the attribute and image feature distances based on the Kullback-Leibler divergence.^23^ FairDist was trained using the Progression Prediction Dataset which contains OCT scans with glaucomatous status and progression outcomes. 70% and 30% of samples were used for model training and testing, respectively.

#### Comparative Methods, Evaluation Metrics and Statistical Analysis

We evaluated the progression prediction performance and demographic equity of five popular deep learning models for processing medical images, including VGG,^24^ DenseNet,^25^ ResNet,^26^ Vision Transformer (ViT),^27^ and EfficientNet.^28^ In addition, we also compared with adversarial training approach to mitigate model biases.^29^ All deep learning modeling and statistical analyses were performed in Python 3.8 (available at http://www.python.org) on a Linux system. Model performance was assessed using the area under the receiver operating characteristic curve (AUC), Sensitivity and Specificity. To assess demographic equity, we adopted the equity-scaled AUC (ES-AUC),^18^ defined as: ***AUC***_***ES***_ = ***AUC***_***overall***_**/(1** + **Σ *AUC***_***overall***_ − ***AUC***_***group***_ ). This metric balances overall AUC with performance disparities among different groups. Statistical significance was evaluated using t-tests and bootstrapping to compare AUC and ES-AUC values between models with and without FAS. Bootstrapping provided confidence intervals and standard error estimates. Results with p < 0.05 were considered statistically significant.

## Results

### Glaucoma Detection Results

We integrated an equity-aware feature attention learning layer into EfficientNet to develop the EqEffNet model. For gender groups, EqEffNet improved the overall AUC by 0.01 (p < 0.05), with the Male group improved from 0.83 to 0.85 (**Figure 3**). For racial groups, both overall AUC and ES-AUC of EqEffNet had an improvement of 0.02 (p < 0.01) compared with EfficientNet, which was contributed by a significant improvement of 0.04 in AUC for White group, although Asians and Blacks had no improvements (**Figure 3**).

**Figure 3:**
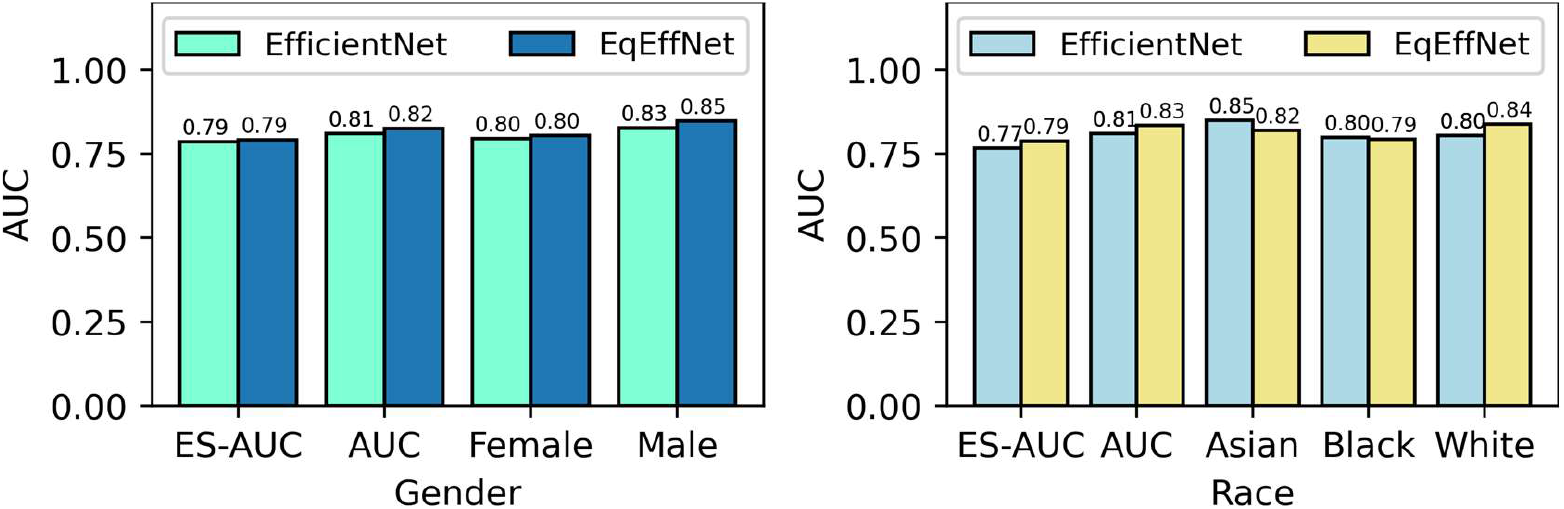
The comparison between EfficientNet and EqEffNet for glaucoma detection. EqEffNet extends EfficientNet by integrating an equity-aware attention mechanism.

### MD Fast Progression Prediction Results

Among the five comparative methods (VGG, DenseNet, ResNet, ViT and EfficientNet) without unfairness mitigation strategies (**Figure 4a**), ResNet achieved the highest overall AUC of 0.69 and ES-AUC of 0.67 (p < 0.05) for the gender attribute, outperforming VGG (0.65 and 0.63), DenseNet (0.68 and 0.65), ViT (0.65 and 0.61), and EfficientNet (0.66 and 0.63). EfficientNet achieved the highest sensitivity of 0.67 (p < 0.01), while both VGG and ResNet demonstrated the highest specificity of 0.66 (p < 0.05) among the five methods (**Figure 4a**). For gender groups, FairDist outperformed all five methods without unfairness mitigation strategies, achieving the highest overall AUC of 0.74 and ES-AUC of 0.69 (p < 0.01). The group AUCs for Females (0.75) and Males (0.79) were consistently higher than those of other methods. Additionally, FairDist matched EfficientNet with the highest sensitivity of 0.67 (**Figure 4a**). While comparing with the two methods with unfairness mitigation on gender groups (**Figure 4b**), FairDist improved the overall AUC, ES-AUC and sensitivity by 0.06 0.11 and 0.20 over EfficientNet + Adversarial (p < 0.01), and 0.05, 0.15 and 0.27 over EqEffNet (p < 0.01), respectively.

**Figure 4:**
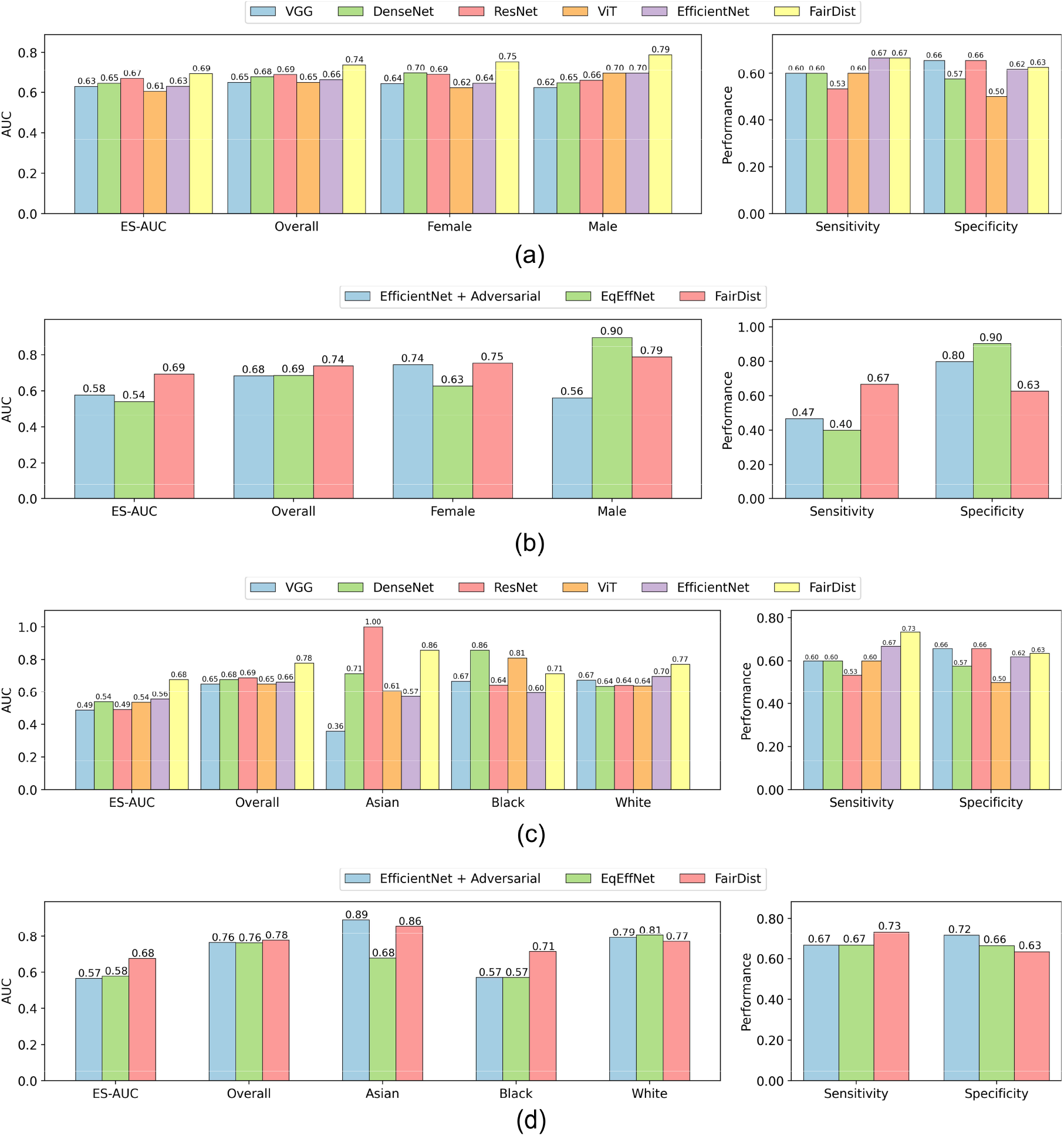
Prediction performance of MD Fast progression. (a) Comparison of baseline methods and the proposed FairDist model for gender groups. (b) Comparison of methods with unfairness mitigation strategies for gender groups.(c) Comparison of baseline methods and the proposed FairDist model for racial groups. (d) Comparison of methods with unfairness mitigation strategies for racial groups. MD: visual field mean deviation. Adversarial: with adversarial training.

For racial groups, ResNet remained the highest AUC of 0.69 among the five baseline methods without fairness considerations. However, its ES-AUC (0.49) and sensitivity (0.53) were lower than those of EfficientNet, which achieved an ES-AUC of 0.56 and sensitivity of 0.67 (**Figure 4c**). FairDist achieved the highest AUC of 0.78, ES-AUC of 0.68 and sensitivity of 0.73 when compared with other methods with and without applying unfairness mitigation strategies (**Figure 4c**). Compared with EqEffNet, FairDist improved the AUCs for Asians and Blacks by 0.20 and 0.14 (p < 0.01), respectively. While comparing with EfficientNet + Adversarial, FairDist improved the AUC in Black group by 0.14 (p < 0.01), although the corresponding AUCs for Asians and Whites had slight drops (**Figure 4d**).

### TD Pointwise Progression Prediction Results

On the gender attribute, EfficientNet achieved an overall AUC of 0.72, ES-AUC of 0.70 and sensitivity of 0.71, which outperformed (p < 0.05) other four methods including VGG, DenseNet (except for sensitivity), ResNet and ViT without designing fairness learning components (**Figure 5a**). FairDist achieved the highest AUC (0.74) and ES-AUC (0.72) compared with all five methods without fairness considerations (**Figure 5a**), and meanwhile outperformed EfficientNet + Adversarial (0.71 and 0.62) and EqEffNet (0.72 and 0.66) with fairness considerations (**Figure 5b**). In addition, FairDist achieved the highest averaged sensitivity and specificity which was 0.67 (**Figure 5a** and **Figure 5b**), compared with VGG (0.60), ResNet (0.60), ViT (0.56), EfficientNet (0.65), EfficientNet + Adversarial (0.66) and EqEffNet (0.62), respectively. FairDist achieved the same specificity of 0.70 as DenseNet, but lower sensitivity (0.64) than DenseNet (0.71).

**Figure 5:**
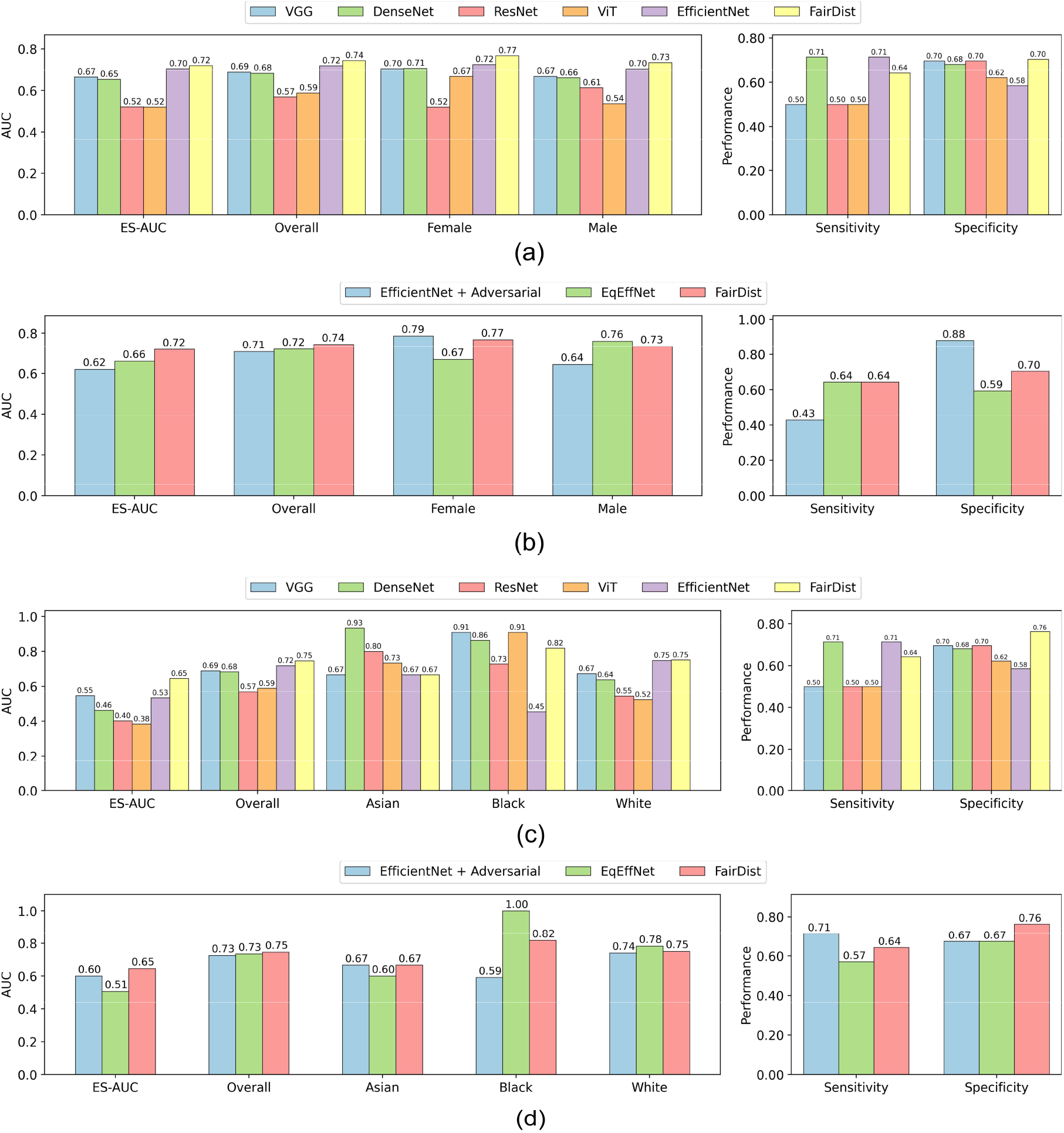
Prediction performance of TD Pointwise progression. (a) Comparison of baseline methods and the proposed FairDist model for gender groups. (b) Comparison of methods with unfairness mitigation strategies for gender groups.(c) Comparison of baseline methods and the proposed FairDist model for racial groups. (d) Comparison of methods with unfairness mitigation strategies for racial groups. MD: visual field mean deviation. Adversarial: with adversarial training.

On the race attribute, FairDist achieved the highest AUC (0.75), ES-AUC (0.65) and specificity (0.76) compared with all other methods (p < 0.05, **Figure 5c** and **Figure 5d**). Although EqEffNet achieved the perfect AUC of 1.0 for Blacks and better AUC than FairDist in White group (**Figure 5d**), its AUC in Asian groups is significantly lower than FairDist which had caused much lower ES-AUC (0.51) than that of FairDist (0.65). FairDist also improved the sensitivity and specificity by 0.07 and 0.09 (p < 0.01), respectively, compared with EqEffNet (**Figure 5d**).

## Discussion

Glaucomatous optic neuropathy characterized by progressive vision loss is the leading cause of irreversible blindness globally.^30^ Currently, there is no effective cure for glaucoma, making it crucial to predict the potential progression of visual field impairment at an early stage. However, glaucoma progression prediction remains underexplored compared to glaucoma detection with deep learning, primarily due to the scarcity of available data.^31^ This challenge is further complicated by deep learning models which have raised significant concerns regarding fairness.^17^ We proposed solutions to address these limitations. These include an equity-aware feature attention learning mechanism and knowledge distillation, which leveraged an equity-enhanced glaucoma detection model trained on substantial samples to improve both performance and demographic equity in progression prediction.

The superiority of the proposed approach stems from two key factors. First, the EqEffNet model, equipped with an equity-aware feature attention mechanism, enhances both performance and equity, as demonstrated in glaucoma detection (**Figure 3**) across gender and racial attributes. The equity-aware attention layer enables the model to adjust feature importance in OCT scans according to the demographic attributes (**Figure 6a**), thus making the model has improved capacity to balance the feature learning of various groups. Additionally, EqEffNet generally outperformed methods lacking unfairness mitigation strategies in progression prediction, showing notable improvements for race in MD Fast progression (**Figure 4c** and **Figure 4d**) and for both gender and race in TD Pointwise progression (Figure 5). Second, the pretrained detection model is beneficial to enhance the glaucoma progression prediction model through knowledge distillation which can be observed by the comparisons between FairDist and EqEffNet (**Figure 4** and **Figure 5**). This allows an equity-enhanced and well-performing glaucoma detection model to guide the feature learning of a progression prediction model, which is beneficial especially when data scarcity is a challenge to train a good model. The averaged gradient-weighted class activation map of FairDist (**Figure 6b**) was visually closed to that of EqEffNet (**Figure 6a**) also exemplified that knowledge distillation has reshaped the progression model to learn features from OCT scans for enhanced model performance and equity. This is also meaningful that features on the OCT retinal layers were more important, which were better captured by FairDist than the baseline EfficientNet model (**Figure 6c**).

**Figure 6:**
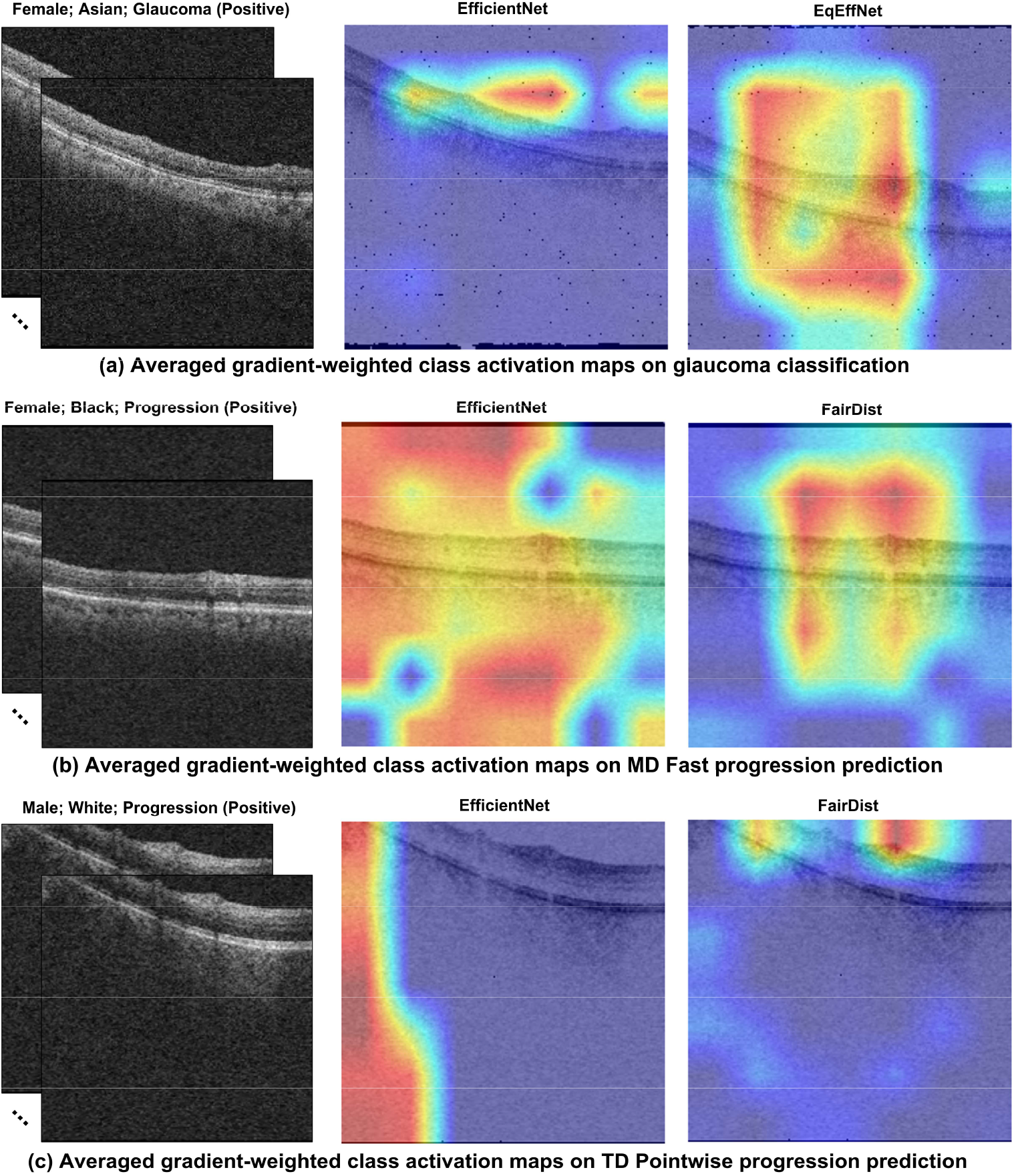
Averaged gradient-weighted class activation maps across all OCT scans. (a) EfficientNet versus EqEffNet on glaucoma classification. (b) EfficientNet versus FairDist on MD Fast progression prediction. (c) EfficientNet versus FairDist on TD Pointwise progression prediction.

Although FairDist consistently achieved highest AUC and ES-AUC scores, compared with other methods with and without unfairness mitigation strategies, it did not always improve the AUCs across different groups. For example, FairDist achieved the best performance of 0.75 (p < 0.05) in Female group among all methods but its AUC (0.79) was worse than that (0.90) of EqEffNet Male group in the prediction of MD Fast progression (**Figure 4b**). For TD Pointwise progression, FairDist did not excel in both Female (AUC: 0.77) and Male (AUC: 0.73) groups, where EfficientNet + Adversarial led the Female (AUC: 0.79) and EqEffNet led the Male (AUC: 0.76) groups, respectively (**Figure 5b**). This is because that mitigating the unfairness of deep learning models is a multi-objective optimization process which aims to reduce group disparities without compromising the overall system performance.^32,33^ This process will inevitably rebalance different groups, leading to performance increase or decrease in certain groups. Therefore, it is meaningful to pursue globally enhanced and equitable model performance across various demographic groups instead of emphasizing certain groups. This can be quantified by the ES-AUC which has coordinated the overall performance and model equity. The proposed FairDist model promises to achieve both improved overall AUC and ES-AUC, as evidenced by the results on gender and racial groups (**Figure 4** and **Figure 5**).

However, the proposed equitable model and its evaluations have several limitations. First, although we studied a critical task of glaucoma progression prediction and meanwhile evaluated the model for glaucoma detection, we have not evaluated the model performance and equity for other disease progression tasks due to a lack of public datasets with demographics. Second, we primarily used the equity-scaled AUC to assess the model equity, while other metrics such as demographic parity and equalized odds.^34,35^ However, it is impossible to satisfy different metrics due to varying definitions and constrains especially in situations of highly-imbalanced distributions of demographics and labels.^36^ Third, we primarily focused on the EfficientNet model to design the equitable deep learning model since it generally outperformed other baseline models for progression predictions in this work. However, it is meaningful to explore more powerful model architectures and training paradigm such as adapting pretrained foundation models to boost the model performance. Forth, we designed an equity-aware attention layer which was helpful to differentiate image feature importance for improved model performance and equity. However, future work necessitates a comprehensive comparison of different unfairness mitigation strategies and compare their generalizability to various demographic groups.^15^ Last, the progression prediction dataset included in this evaluation is not substantial, which is a critical challenge that hinders the advancement of develop equitable deep learning models. Further work will test the generality of the proposed model using an increased scale of dataset.

In conclusion, we proposed an equity-aware deep learning model with knowledge distillation to enhance the model performance and equity of glaucoma progression prediction. The model has been tested on MD Fast and TD Pointwise progressions, achieving improved performance compared with methods with and without fairness learning components. Our approach has the potential to improve demographic equity in glaucoma progression and it can also be generalized to other disease progression prediction tasks.

## Data Availability

All data produced are available online at https://github.com/Harvard-Ophthalmology-AI-Lab/Harvard-GDP

## Notes

### Competing Interest Statement

The authors have declared no competing interest.

### Funding Statement

This study did not receive any funding

### Author Declarations

Two publicly accessible datasets were used to develop deep learning models. The datasets used in this study were fully de-identified. This study complied with the guidelines outlined in the Declaration of Helsinki. In light of the study's retrospective design, the requirement for informed consent was waived.

## References

1. Quigley HA. The number of people with glaucoma worldwide in 2010 and 2020. British Journal of Ophthalmology. 2006;90(3):262–267. doi:10.1136/bjo.2005.081224

2. Friedman DS, Wolfs RCW, O’Colmain BJ, et al. Prevalence of open-angle glaucoma among adults in the United States. Arch Ophthalmol. 2004;122(4):532–538.

3. Jayaram H, Kolko M, Friedman DS, Gazzard G. Glaucoma: now and beyond. The Lancet. 2023;402(10414):1788–1801. doi:10.1016/S0140-6736(23)01289-8

4. Thompson AC, Jammal AA, Medeiros FA. A Review of Deep Learning for Screening, Diagnosis, and Detection of Glaucoma Progression. Transl Vis Sci Technol. 2020;9(2):42. doi:10.1167/tvst.9.2.42

5. Zhang X, Dastiridou A, Francis BA, et al. Comparison of Glaucoma Progression Detection by Optical Coherence Tomography and Visual Field. Am J Ophthalmol. 2017;184:63–74. doi:10.1016/j.ajo.2017.09.020

6. Hou K, Bradley C, Herbert P, et al. Predicting Visual Field Worsening with Longitudinal OCT Data Using a Gated Transformer Network. Ophthalmology. 2023;130(8):854–862. doi:10.1016/j.ophtha.2023.03.019

7. Mohammadzadeh V, Wu S, Besharati S, et al. Prediction of Visual Field Progression with Baseline and Longitudinal Structural Measurements Using Deep Learning. Am J Ophthalmol. 2024;262:141–152. doi:10.1016/j.ajo.2024.02.007

8. Mariottoni EB, Datta S, Shigueoka LS, et al. Deep Learning–Assisted Detection of Glaucoma Progression in Spectral-Domain OCT. Ophthalmol Glaucoma. 2023;6(3):228–238. doi:10.1016/j.ogla.2022.11.004

9. Shi M, Tian Y, Luo Y, Elze T, Wang M. RNFLT2Vec: Artifact-corrected representation learning for retinal nerve fiber layer thickness maps. Med Image Anal. 2024;94:103110. doi:10.1016/j.media.2024.103110

10. Min Shi, Jessica A. Sun, Anagha Lokhande, et al. Artifact Correction in Retinal Nerve Fiber Layer Thickness Maps Using Deep Learning and Its Clinical Utility in Glaucoma. Transl Vis Sci Technol. Published online 2023.

11. Li F, Wang D, Yang Z, et al. The AI revolution in glaucoma: Bridging challenges with opportunities. Prog Retin Eye Res. 2024;103:101291. doi:10.1016/j.preteyeres.2024.101291

12. Huang X, Islam MR, Akter S, et al. Artificial intelligence in glaucoma: opportunities, challenges, and future directions. Biomed Eng Online. 2023;22(1):126. doi:10.1186/s12938-023-01187-8

13. Shon K, Sung KR, Shin JW. Can Artificial Intelligence Predict Glaucomatous Visual Field Progression? A Spatial–Ordinal Convolutional Neural Network Model. Am J Ophthalmol. 2022;233:124–134. doi:10.1016/j.ajo.2021.06.025

14. Thakur A, Goldbaum M, Yousefi S. Predicting Glaucoma before Onset Using Deep Learning. Ophthalmol Glaucoma. 2020;3(4):262–268. doi:10.1016/j.ogla.2020.04.012

15. Caton S, Haas C. Fairness in Machine Learning: A Survey. ACM Comput Surv. 2024;56(7):1–38. doi:10.1145/3616865

16. Jin R, Xu Z, Zhong Y, et al. FairMedFM: Fairness Benchmarking for Medical Imaging Foundation Models. arXiv preprint arXiv:240700983. Published online 2024.

17. Chen RJ, Wang JJ, Williamson DFK, et al. Algorithmic fairness in artificial intelligence for medicine and healthcare. Nat Biomed Eng. 2023;7(6):719–742. doi:10.1038/s41551-023-01056-8

18. Luo Y, Tian Y, Shi M, et al. Harvard Glaucoma Fairness: A Retinal Nerve Disease Dataset for Fairness Learning and Fair Identity Normalization. IEEE Trans Med Imaging. 2024;43(7):2623–2633. doi:10.1109/TMI.2024.3377552

19. Tian Y, Shi M, Luo Y, Kouhana A, Elze T, Wang M. FairSeg: A Large-Scale Medical Image Segmentation Dataset for Fairness Learning Using Segment Anything Model with Fair Error-Bound Scaling. In: The Twelfth International Conference on Learning Representations.

20. Xu Z, Li J, Yao Q, Li H, Zhao M, Zhou SK. Addressing fairness issues in deep learning-based medical image analysis: a systematic review. NPJ Digit Med. 2024;7(1):286. doi:10.1038/s41746-024-01276-5

21. Wang M, Tichelaar J, Pasquale LR, et al. Characterization of Central Visual Field Loss in End-stage Glaucoma by Unsupervised Artificial Intelligence. JAMA Ophthalmol. 2020;138(2):190. doi:10.1001/jamaophthalmol.2019.5413

22. Vesti E, Johnson CA, Chauhan BC. Comparison of Different Methods for Detecting Glaucomatous Visual Field Progression. Investigative Opthalmology & Visual Science. 2003;44(9):3873. doi:10.1167/iovs.02-1171

23. Zhang Y, Pan J, Li LK, et al. On the properties of Kullback-Leibler divergence between multivariate Gaussian distributions. Adv Neural Inf Process Syst. 2024;36.

24. Simonyan K, Zisserman A. Very Deep Convolutional Networks for Large-Scale Image Recognition. Published online September 4, 2014.

25. Huang G, Liu Z, Van Der Maaten L, Weinberger KQ. Densely Connected Convolutional Networks. In: 2017 IEEE Conference on Computer Vision and Pattern Recognition (CVPR). IEEE; 2017:2261–2269. doi:10.1109/CVPR.2017.243

26. He K, Xiangyu Zhang, Shaoqing Ren, Jian Sun. Deep residual learning for image recognition. In: IEEE Conference on Computer Vision and Pattern Recognition. ; 2016:770–778.

27. Dosovitskiy A, Beyer L, Kolesnikov A, et al. An Image is Worth 16×16 Words: Transformers for Image Recognition at Scale. Published online October 22, 2020.

28. Tan M, Le Q V. EfficientNet: Rethinking Model Scaling for Convolutional Neural Networks. Published online May 28, 2019.

29. Yang J, Soltan AAS, Eyre DW, Yang Y, Clifton DA. An adversarial training framework for mitigating algorithmic biases in clinical machine learning. NPJ Digit Med. 2023;6(1):55. doi:10.1038/s41746-023-00805-y

30. Tham YC, Li X, Wong TY, Quigley HA, Aung T, Cheng CY. Global prevalence of glaucoma and projections of glaucoma burden through 2040: a systematic review and meta-analysis. Ophthalmology. 2014;121(11):2081–2090.

31. Li J, Cairns BJ, Li J, Zhu T. Generating synthetic mixed-type longitudinal electronic health records for artificial intelligent applications. NPJ Digit Med. 2023;6(1):98. doi:10.1038/s41746-023-00834-7

32. Luo Y, Shi M, Khan MO, et al. FairCLIP: Harnessing Fairness in Vision-Language Learning. In: Proceedings of the IEEE/CVF Conference on Computer Vision and Pattern Recognition (CVPR). ; 2024:12289–12301.

33. Tian Y, Wen C, Shi M, et al. FairDomain: Achieving Fairness in Cross-Domain Medical Image Segmentation and Classification. arXiv preprint arXiv:240708813. Published online 2024.

34. Hardt M, Price E, Srebro N. Equality of opportunity in supervised learning. Adv Neural Inf Process Syst. 2016;29.

35. Mehrabi N, Morstatter F, Saxena N, Lerman K, Galstyan A. A Survey on Bias and Fairness in Machine Learning. ACM Comput Surv. 2022;54(6):1–35. doi:10.1145/3457607

36. Kleinberg J, Mullainathan S, Raghavan M. Inherent trade-offs in the fair determination of risk scores. arXiv preprint arXiv:160905807. Published online 2016.

